# Assessing the adherence of intervertebral disc degeneration clinical trial protocols to SPIRIT guidelines

**DOI:** 10.1101/2024.10.17.24315674

**Authors:** Francis Chemorion, Marc-Antonio Bisotti

## Abstract

**Introduction:** Clinical trials are important for advancing medical knowledge and protocols are the core documents that facilitate their appraisal. The SPIRIT (Standard Protocol Items: Recommendations for Interventional Trials) checklist offers a framework for standardizing the quality and content of clinical trial protocols. Although there is a high growth in number of clinical trials investigating the efficacy and safety of various therapies, many still exhibit deficiencies in information contained in both their reports and protocols. The main objective for this study were to check the findability of intervertebral disc degeneration (IVD) clinical trial protocols and assessing their adherence to SPIRIT recommendations.

**Methods:** We included randomized controlled trials (RCTs) that investigated various therapeutic interventions for IVD provided they included a control group and reported at least one relevant clinical outcome. Studies were excluded if they were not recruiting, were observational, case reports, reviews, meta-analyses, editorials, or commentaries, or if they lacked complete data, such as missing results or a protocol. A search was conducted on ClinicalTrials.gov using terms related to IVD, covering the period from January 2013 to the present, aligning with the post-SPIRIT recommendation publication period. Data extraction was performed using a reduced SPIRIT checklist to assess adherence to protocol guidelines, with compliance measured across 64 key items. A narrative synthesis was conducted to summarize study characteristics, adherence levels and patterns by intervention and sponsor type.

**Results:** Adherence rates vary from 28.13% to 98.44% with a median of 48.44%. Heatmaps revealed heterogeneity, highlighting areas of consistent adherence and regions requiring improvement. High adherence was noted in inclusion criteria and outcome measurement, while lower in research ethics and funding declarations. Industry-sponsored studies demonstrated highest adherence in ‘DRUG’ (79.69%) and ‘DEVICE’ (62.50%) categories, while non-industry sponsors were most adherent in ‘BIOLOGICAL’ (50.00%) and ‘OTHER’ intervention categories.

**Discussion:** Our findings shows a critical need for findability of clinical trial protocols to assist in appraisal of interventions and enhanced adherence to SPIRIT guidelines among IVD clinical trials, for greater transparency and accuracy. Adherence is essential for ensuring high-quality evidence that can inform clinical practice and patient care in managing IVD.

## 1 Introduction

### 1.1 Background

Clinical trial protocols directly facilitate the generation of primary evidence on treatment efficacy, effectiveness, and safety in medical research (Butcher et al., 2020). These documents serve as the blueprint for conducting clinical trials, outlining the study’s objectives, design, methodology, statistical considerations, and organization (Chan & Hróbjartsson, 2018). The publication and accessibility of these protocols is crucial for mitigation of selective reporting (Whittington, 2018), Enhancement of internal validity (Chan & Hróbjartsson, 2018), Reduction of statistical bias (Gryaznov et al., 2020) and Increased reproducibility (Cuschieri, 2019). In cases where novel experimental approaches are being employed, the publication of protocols is particularly beneficial, as it allows for public scrutiny and peer review, especially in situations with high potential for bias (Whittington, 2018).

### 1.2 Components and Importance of Clinical Trial Protocols

Clinical trial protocols include essential elements such as study background, rationale, methodological procedures, relevance to clinical practice, administrative details, and ethical safeguards (An Wen Chan et al., 2013). Public availability of these protocols enhances scientific integrity by preventing research falsification, increasing the visibility of randomized controlled trials (RCTs), and improving the overall quality and reliability of their results (Gryaznov et al., 2020). Transparency also facilitates peer review, leading to better study design and execution (Taichman et al., 2017). Conversely, low-quality protocols lacking key elements can compromise the integrity of an RCT, resulting in unreliable data and affecting the validity of its conclusions (Gryaznov et al., 2020; Jennifer Tetzlaff et al., 2012). Therefore, creating and disseminating high-quality protocols is crucial for ensuring the accuracy and reproducibility of clinical research.

### 1.3 The SPIRIT Guidelines

In response to the need for standardization and quality improvement in clinical trial protocols, the Standard Protocol Items: Recommendations for Interventional Trials (SPIRIT) guidelines were published in 2013 (An Wen Chan et al., 2013). These guidelines offer a comprehensive 33-item checklist aimed at enhancing the completeness and quality of clinical trial protocols. The adoption of SPIRIT guidelines has proven pivotal in addressing critical aspects of trial design, execution, and reporting, thereby facilitating research reproducibility and outcome reliability (Cuschieri, 2019).

Key areas addressed by the SPIRIT guidelines include:

### 1.4 Intervertebral Disc Degeneration (IVD) and Clinical Trials

Intervertebral disc degeneration (IVD) represents a significant health concern with substantial impact on quality of life and healthcare systems globally (Hartvigsen et al., 2018). As the prevalence of IVD continues to grow, there has been a corresponding increase in clinical trials investigating various therapeutic interventions (Maher et al., 2017). These interventions span a wide range, including biological therapies such as stem cell treatments and growth factors, medical devices like artificial discs and minimally invasive surgical tools, pharmacological interventions including anti-inflammatory drugs and pain modulators, and physical therapies and rehabilitation approaches (AO Research Institute Davos, Clavadelerstrasse 8, 7270 Davos, Switzerland et al., 2013; Samartzis et al., 2013). The diversity and complexity of these interventions highlight the critical importance of maintaining high-quality, transparent clinical trial protocols in IVD research. As noted by (Yim et al., 2014), the rapidly evolving landscape of IVD treatments necessitates thorough methodological standards to ensure the validity and reliability of clinical findings, thereby facilitating evidence-based decision-making in patient care and policy development.

### 1.5 Knowledge Gap and Study Rationale

Despite the recognized importance of protocol quality and adherence to SPIRIT guidelines in elevating research standards, there remains a significant gap in the literature regarding the availability and quality of clinical trial protocols for specific medical conditions, including IVD. Investigations into the accessibility of IVD clinical trial protocols and their adherence to SPIRIT recommendations have not been comprehensively conducted. This gap limits the ability to critically evaluate and enhance the reporting quality of interventions within this domain (Chan & Hróbjartsson, 2018). By assessing the availability and quality of IVD clinical trial protocols, we can identify areas of strength and weakness in current practices, paving the way for targeted improvements in trial design and reporting within this critical area of medical research.

### 1.6 Study Objectives

The primary objectives of this study are:

1. To assess the findability and accessibility of IVD clinical trial protocols, cataloguing their availability to the research community and the general public.
2. To evaluate the adherence of available IVD clinical trial protocols to the SPIRIT guidelines, involving a detailed assessment of the protocols’ structure, content, and the presence of essential elements as stipulated by the SPIRIT checklist.
3. To compare the quality and completeness of protocols between industry-sponsored and non-industry-sponsored IVD clinical trials protocols.

By addressing these objectives, this study aims to provide valuable insights into the current state of IVD clinical trial reporting and identify opportunities for improving the transparency, reproducibility, and overall quality of research in this important field of medicine.

## 2 Methods

### 2.1 Eligibility Criteria

The following studies were included:

(1) studies that focus on patients diagnosed with intervertebral disc degeneration (IVD);
(2) studies that investigate various forms of treatment for IVD, such as pharmacological, surgical, regenerative, or other therapeutic interventions;
(3) studies that include a control group, which may be standard care, placebo, or an alternative intervention;
(4) studies that report on at least one relevant clinical outcome, such as pain reduction, improvement in function, or quality of life; and
(5) only randomized controlled trials (RCTs) as they provide the highest level of evidence for treatment efficacy.

RCTs that were not yet recruiting, active but not recruiting, terminated, withdrawn, or had an unknown status or those that are observational, case reports, reviews, meta-analyses, editorials, commentaries, and qualitative or that contain incomplete data for example those that lack results or a formal clinical trial protocol or those that are enrolling participants by invitation only or those that have been suspended were excluded.

### 2.2 Information Sources and Search Strategy

A targeted search was conducted on ClinicalTrials.gov for clinical trials related to intervertebral disc degeneration, using relevant search terms such as “intervertebral disc degeneration” and “disc degeneration.” The search parameters were set from January 2013 to the present to align with the post-SPIRIT recommendation publication period.

### 2.3 Data Extraction

The eligible clinical trial protocols were assessed using the SPIRIT checklist, which originally consists of 270 items across 33 sections. To streamline the assessment and focus on elements most pertinent to this study, we reduced the checklist to 64 items. The reduction was guided by the PICOS framework (Population, Intervention, Comparator, Outcome, and Study Design), ensuring that only items directly relevant to defining the research question and evaluating the essential elements of trial design were included (Gryaznov et al., 2020). This approach prioritized checklist items that could significantly impact the clarity, accuracy and transparency of clinical trial reporting. For each of the selected SPIRIT checklist items, a score of 0 was assigned if the item was not found in the protocol, and 1 if it fully complied with the guidelines.

An illustration of the data collection table is as shown in Table 1 below.

**Table 1.** SPIRIT Checklist Key Sections *(An Wen Chan et al*., *2013*)

| Section | Description |
| --- | --- |
| Administrative information | Title, trial registration, protocol version, funding and roles and responsibilities |
| Introduction | Scientific background, explanation of rationale and objectives |
| Methods: Participants, interventions, and outcomes | Study setting, Eligibility criteria, interventions, outcomes, sample size, participant timeline and recruitment |
| Methods: Assignment of interventions (for controlled trials) | Allocation( sequence generation, allocation concealment mechanism, implementation and binding(masking) ) |
| Methods: Data collection, management, and analysis | Data collection methods, data management and statistical methods |
| Methods: Monitoring | Data monitoring, harms and auditing |
| Ethics and dissemination | Research ethics approval, protocol amendments, consent or assent, confidentiality, declaration of interests, access to data, ancillary and post-trial care and dissemination policy |
| Appendices | Informed consent materials and biological specimen |

### 2.4 Data Synthesis

A narrative synthesis was chosen over a quantitative synthesis, such as a meta-analysis, due to the heterogeneity observed in the study protocols, interventions, and adherence levels to the SPIRIT guidelines. The variability in study designs, intervention types, and reporting practices made a quantitative synthesis inappropriate, as pooling data in a meta-analysis would not have accurately represented the diverse characteristics and contexts of the included trials. Instead, the narrative synthesis approach allowed for a comprehensive and structured analysis of the findings, capturing the variations across the studies. The results were summarized in tables and figures, presenting the following:

- Characteristics of included studies
- Adherence to SPIRIT guidelines (by trial and intervention type)
- Most and least adhered to SPIRIT items
- Adherence by category and sponsor type
- Adherence by SPIRIT category

## 3 Results

### 3.1 Study Selection

The search revealed 469 intervertebral disc degeneration (IVD) clinical trials. After an initial screening, trials that were recruiting or not yet recruiting, trials that were actively not recruiting and terminated, trials that were enrolling by invitation and were suspended, and trials that were withdrawn and unknown studies were excluded, 244 studies were selected for closer review and further narrowed to 156 studies after excluding observational studies. The other exclusion criteria that led to the removal of more studies at later stages were 138 for lacking results and protocols. This selection process resulted in 18 trials that spanned from 2013 to 2022.

### 3.2 Study Characteristics

From the selected studies, a wide range of interventions were utilized to address various spinal conditions, particularly degenerative disc disease. Drug-based interventions were featured in 4 studies (24%), indicating a significant interest in pharmacological treatments for pain management and inflammation associated with spinal degeneration. Biological interventions, including cell-based therapies and bone grafts, were examined in 5 studies (29%), underscoring the growing exploration of regenerative medicine to promote spinal repair and regeneration. Device-based interventions, such as artificial discs, spinal cages, and fusion systems, were the most commonly studied, comprising 9 studies (53%), reflecting the substantial focus on innovative surgical and implantable devices to improve clinical outcomes. Additionally, 2 studies (12%) investigated alternative interventions, including behavioural and rehabilitative approaches, showcasing the multidisciplinary strategies being explored to enhance spinal health management.

Table 2 provides a comprehensive overview of these studies, detailing the status of each study (such as completed, terminated, or recruiting), the specific types of interventions being investigated and the sponsorship source (whether industry or non-industry). This table serves as a crucial reference for understanding the characteristics and scope of the selected studies, highlighting the variation in research focus and methodology across different types of interventions.

**Table 2.**
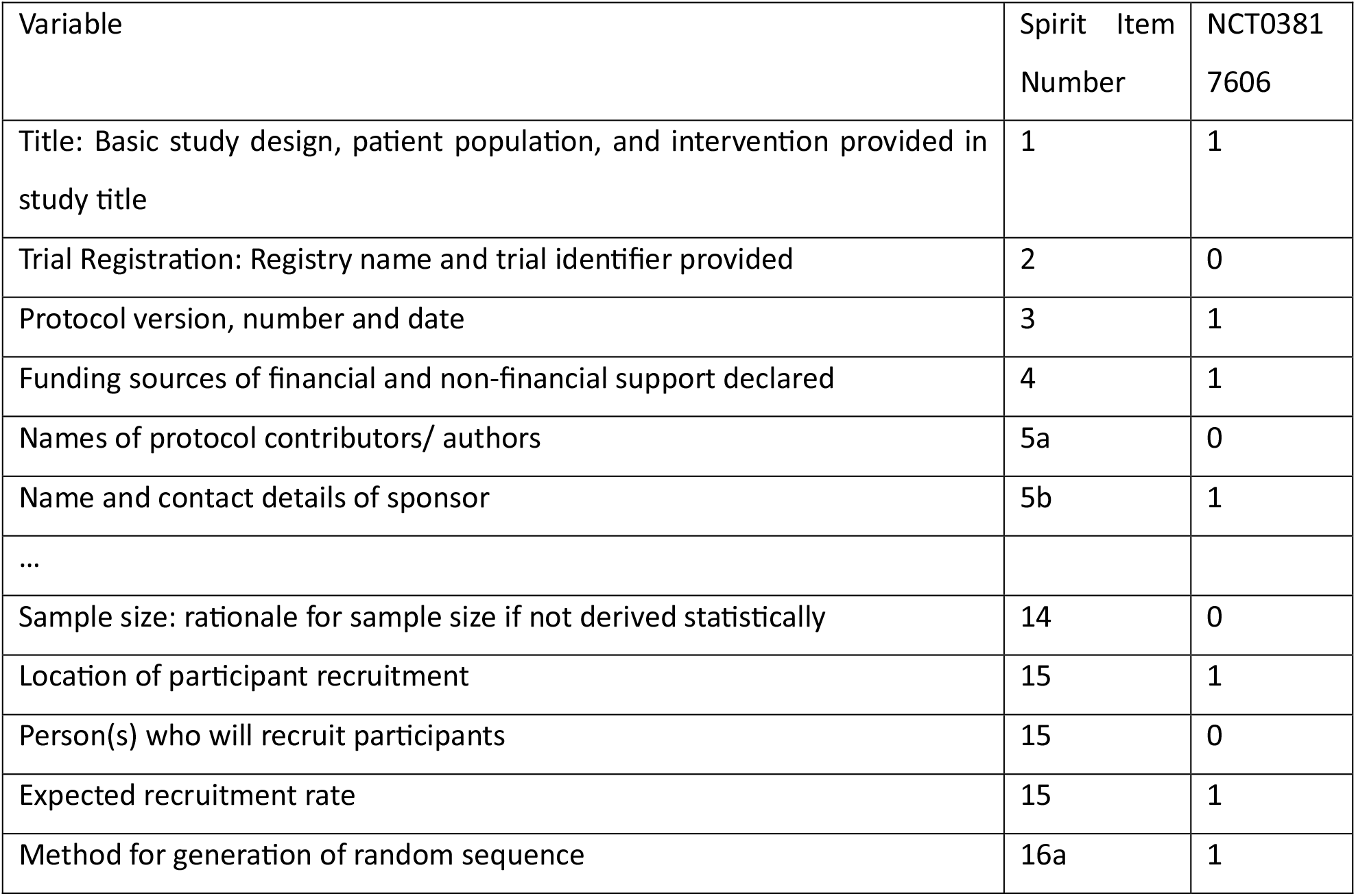
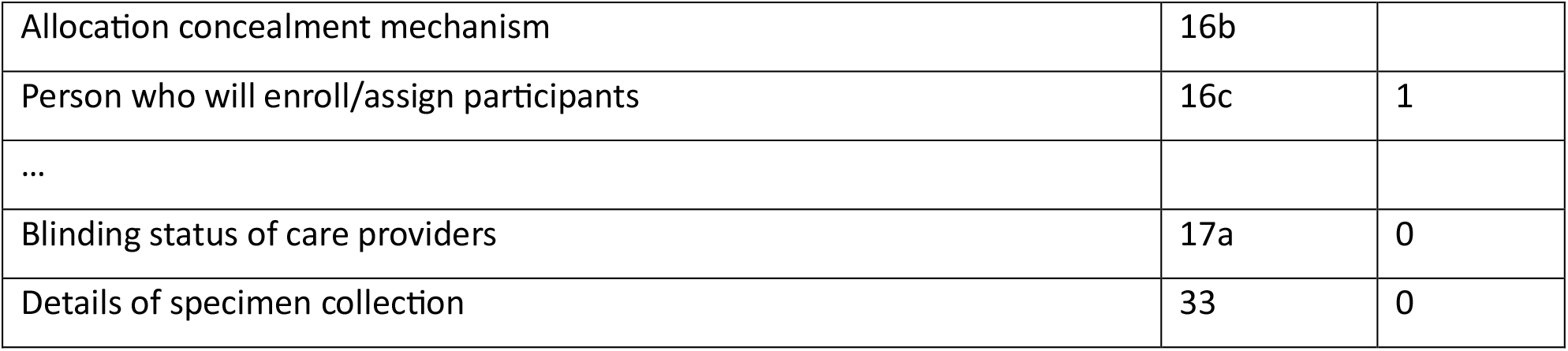
Illustration of Data Extraction according to the SPIRIT checklist.

| Variable | Spirit Item Number | NCT03817606 |
| --- | --- | --- |
| Title: Basic study design, patient population, and intervention provided in study title | 1 | 1 |
| Trial Registration: Registry name and trial identifier provided | 2 | 0 |
| Protocol version, number and date | 3 | 1 |
| Funding sources of financial and non-financial support declared | 4 | 1 |
| Names of protocol contributors/ authors | 5a | 0 |
| Name and contact details of sponsor | 5b | 1 |
| ... |  |  |
| Sample size: rationale for sample size if not derived statistically | 14 | 0 |
| Location of participant recruitment | 15 | 1 |
| Person(s) who will recruit participants | 15 | 0 |
| Expected recruitment rate | 15 | 1 |
| Method for generation of random sequence | 16a | 1 |
| Allocation concealment mechanism | 16b |  |
| Person who will enroll/assign participants | 16c | 1 |
| ... |  |  |
| Blinding status of care providers | 17a | 0 |
| Details of specimen collection | 33 | 0 |

**Table 3.** Characteristics of Selected Study Protocols.

| **Study Status** | **TERMINATED** | **WITHDRAWN** | **COMPLETE** | **UNKNOWN** | **ACTIVE_NOT_RECRUITING** | **Total** |
| --- | --- | --- | --- | --- | --- | --- |
| Sample Size, median (IQR) | 14 (6 - 20) | 0 (0-0) | 61 (23 -115) | 0 (0-0) | 123 (123-123) | 24(20-96) |
| **Interventions** |  |  |  |  |  |  |
| Device | 4 (33.3%) | 0(0%) | 8 (66.7%) | 0(0%) | 0(0%) | 12 |
| Drug | 2 (22%) | 0(0%) | 5 (55.6%) | 0(0%) | 2 (22.2%) | 9 |
| Biological | 0 (0%) | 2 (40%) | 6 (60%) | 0 (0%) | 0 (0%) | 5 |
| Behavioural | 0 (0%) | 0 (0%) | 1 (100%) | 0 (0%) | 1 (100%) | 1 |
| Diagnostic Test | 0 (0%) | 0 (0%) | 0 (0%) | 0 (0%) | 0 (0%) | 1 |
| Other | 1 (33.3%) | 0 (0%) | 2 (66.7%) | 0 (0%) | 0 (0%) | 3 |
| **Sponsorship** |  |  |  |  |  |  |
| Industry | 5 (83.3%) | 0 (0%) | 3 (27.3%) | 0 (0%) | 1 (100%) | 12 |
| Non-industry | 2 (50%) | 0 (0%) | 3 (30%) | 0 (0%) | 0 (0%) | 5 |

## 4 Discussion

### 4.1 Overall Adherence

The overall adherence across the evaluated dataset varied widely among different columns, with compliance percentages ranging from 28.13% to 98.44%. The median compliance was 48.44%, indicating that at least half of the dataset’s columns had a compliance rate above this level. Approximately 25% of the columns showed compliance rates below 43.75% (the 25th percentile), while 25% exhibited compliance rates above 62.5% (the 75th percentile), highlighting areas with significantly lower or higher adherence levels. This variation suggests potential opportunities for targeted improvements in compliance across the dataset.

The binary heatmap below further provides a detailed visualization of adherence patterns across various clinical trial protocols, with dark shaded cells indicating adherence and light shaded cells representing non-adherence. This visualization allows for a quick identification of consistent adherence or non-adherence within and across different studies. The varying density of dark and light shaded cells suggests substantial heterogeneity in adherence rates, reflecting that certain studies or protocols exhibit high levels of adherence, while others show frequent non-adherence. Some clinical trial protocols demonstrate uniform adherence across all items, which could indicate well-structured study protocols or effective implementation strategies. In contrast, other studies show more mixed patterns, with clusters of non-adherence, potentially highlighting specific areas where protocol requirements are challenging to meet or less emphasized. These findings can guide targeted improvements, directing attention to the studies or protocol areas most in need of adherence reinforcement, thereby enhancing overall compliance across the dataset.

### 4.2 Most adhered SPIRIT Recommendations

As observed in Figure 4 below, the top of the heatmap, showing darker shades, indicates the SPIRIT components with the highest adherence. While the data does not show 100% adherence across all types, the components such as “Inclusion and exclusion criteria for trial participants described”, “Primary Outcome: specific measurement variable”, “Primary outcomes: time point of measurement”, and “Who has authority to stop the trial” are among the most adhered to, demonstrating high compliance across various types of sponsorships.

**Figure 1.**
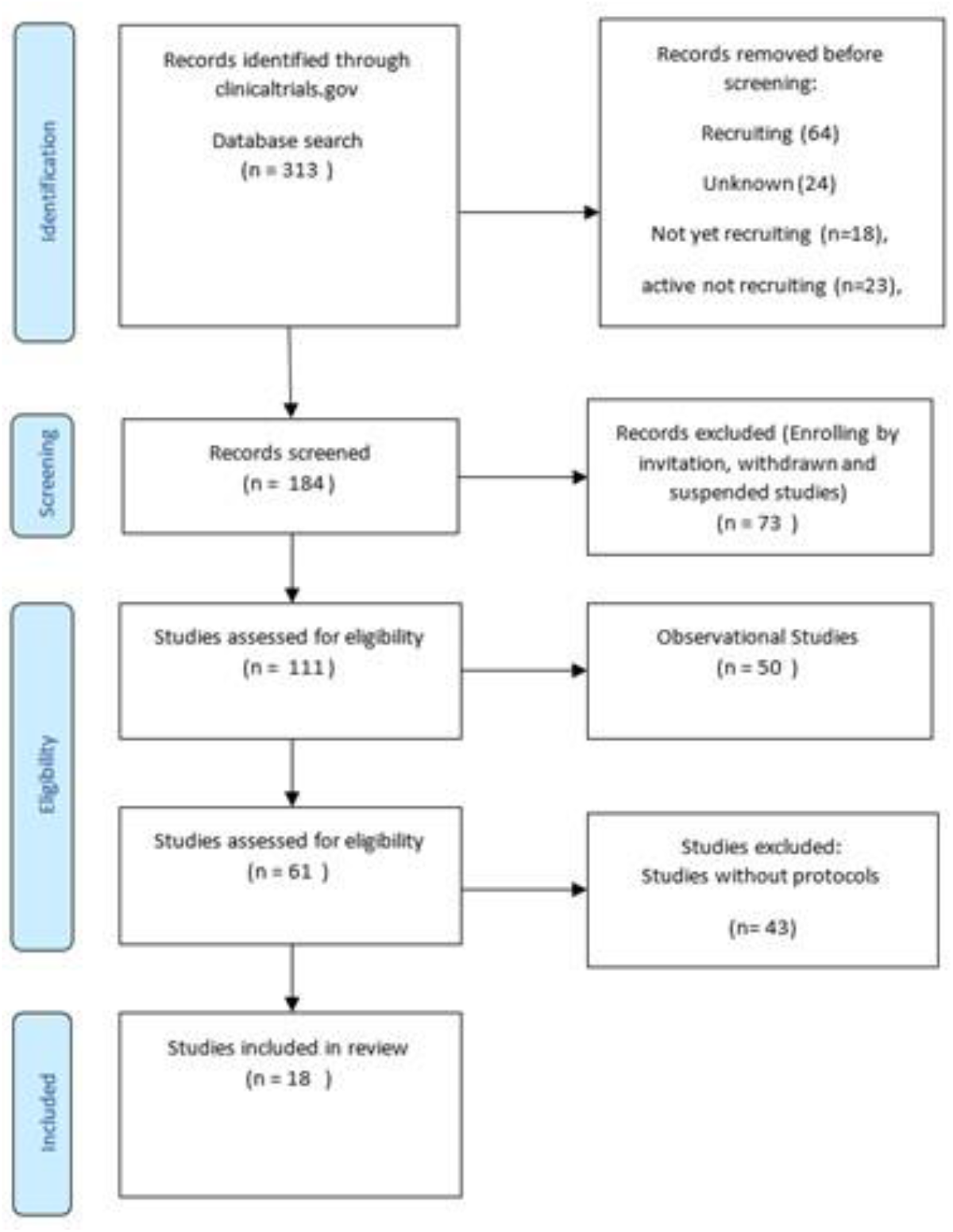
Selection of IVD Clinical Trial Protocols

**Figure 2.**
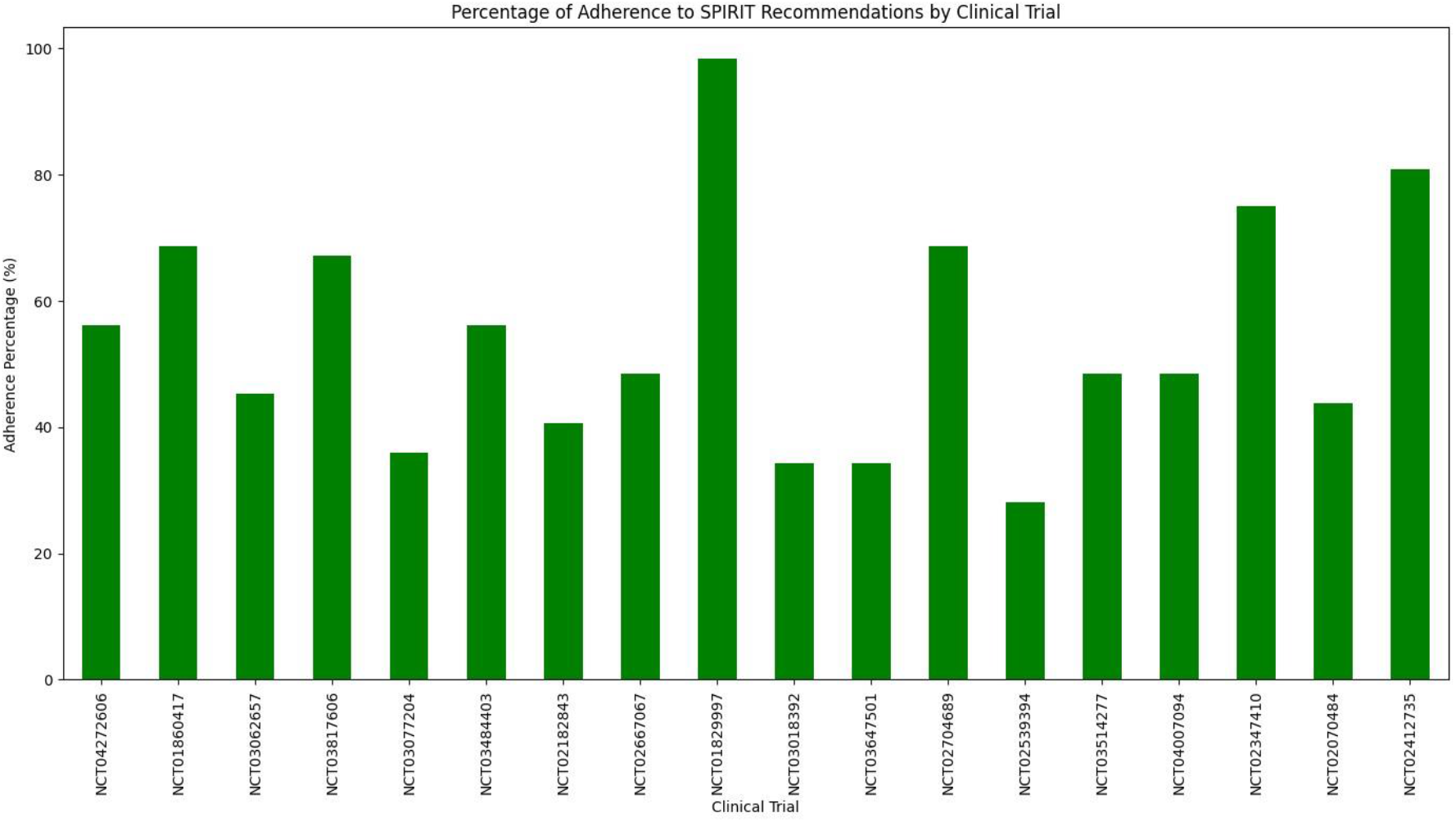
Percentage Adherence by Trial

**Figure 3.**
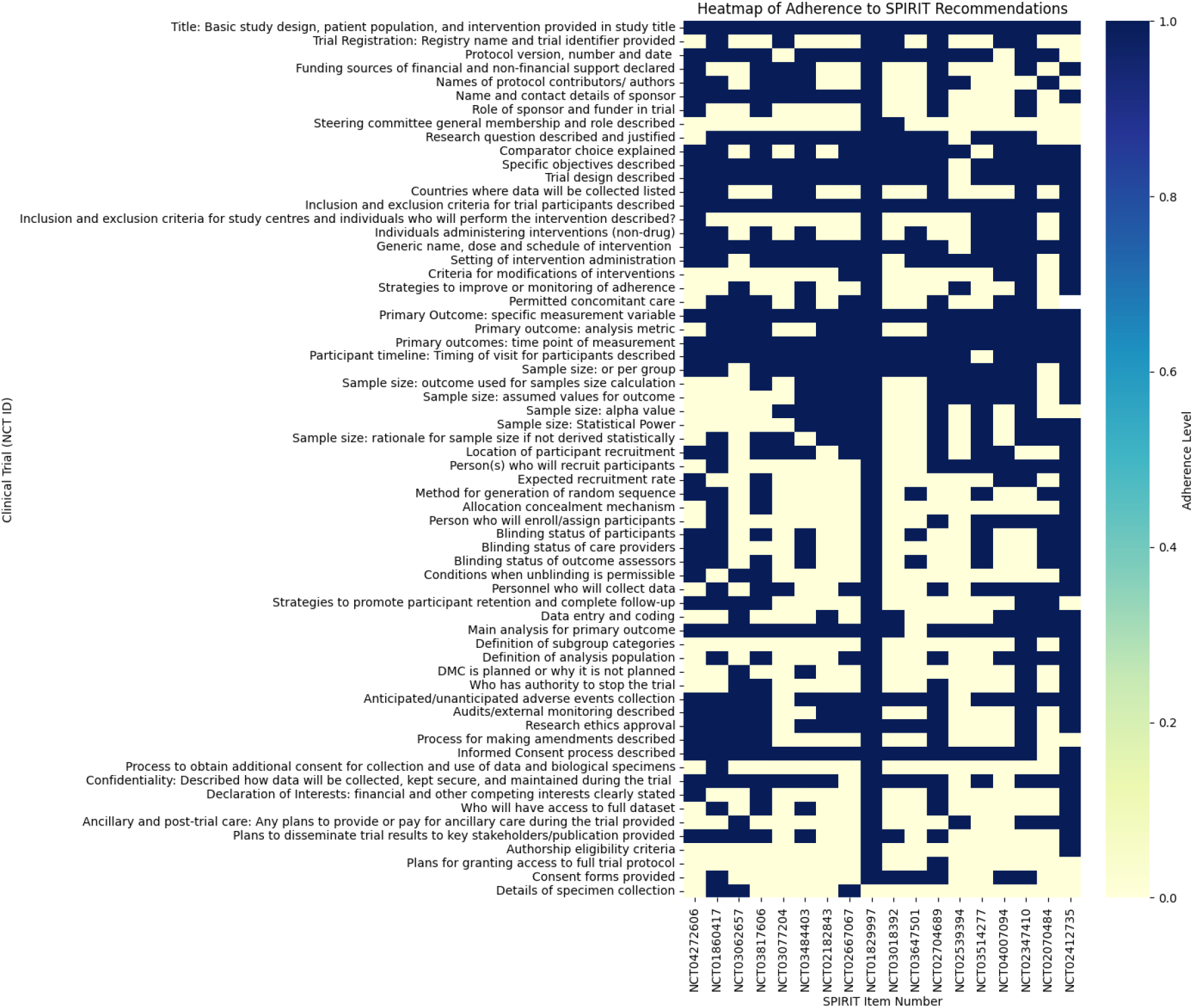
A Heatmap of SPIRIT Adherence

**Figure 4.**
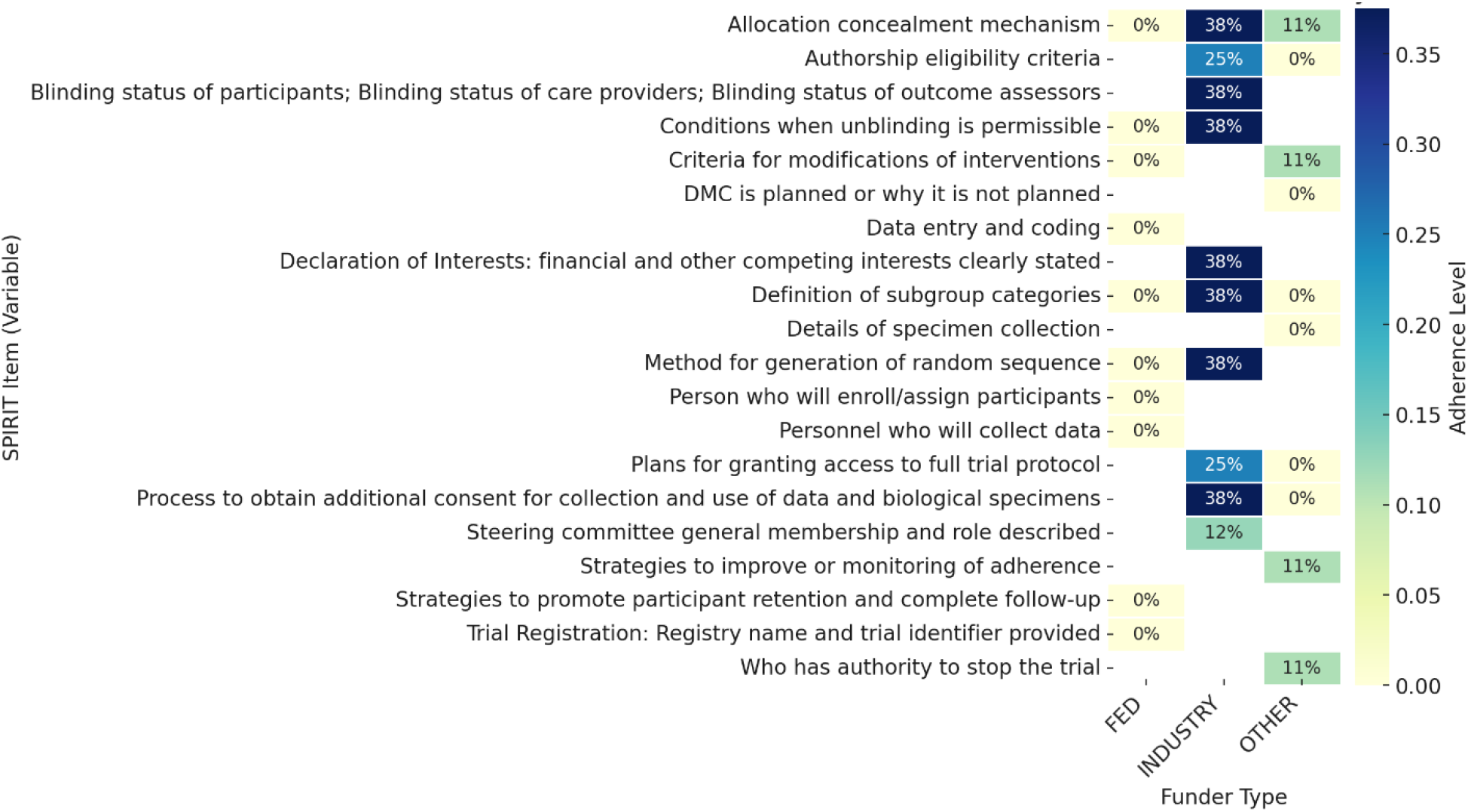
Least Adhered SPIRIT Recommendations

The items with lighter shades, including “Personnel who will collect data” and “Strategies to promote participant retention and complete follow-up”, exhibit room for improvement in terms of adherence, especially in industry-sponsored trials.

### 4.3 Least Adhered SPIRIT Recommendations

As depicted in Figure 5 below, the following SPIRIT components show the lowest adherence levels: “Research ethics approval”, “Protocol version, number and date”, “Consent forms provided”, and “Funding sources of financial and non-financial support declared”. These components have varied adherence levels, with some as low as 40% in industry sponsorship and 50% in non-industry sponsorship.

**Figure 5.**
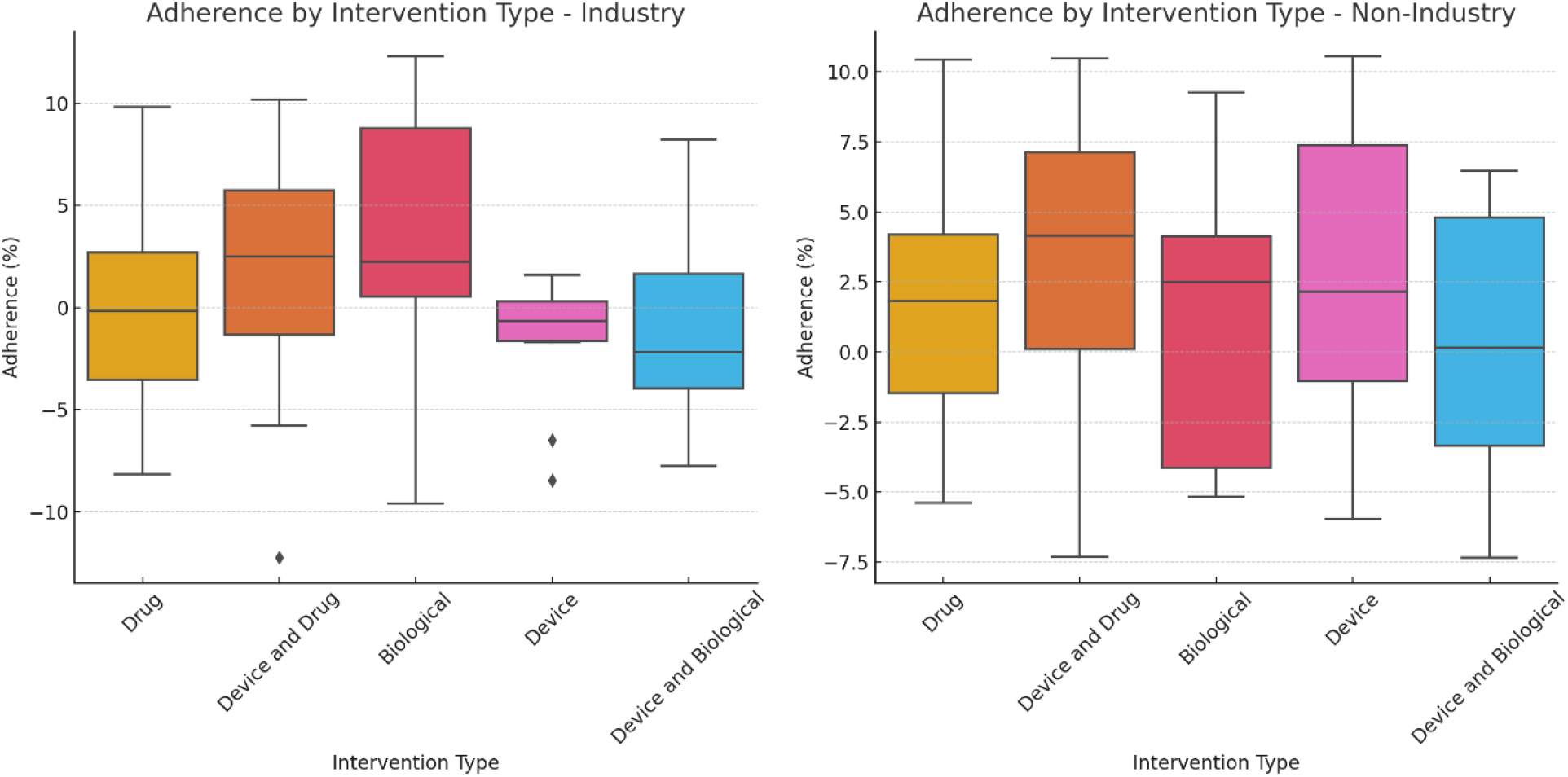
Average Adherence by Intervention Category in Industry vs Non-Industry

**Figure 6.**
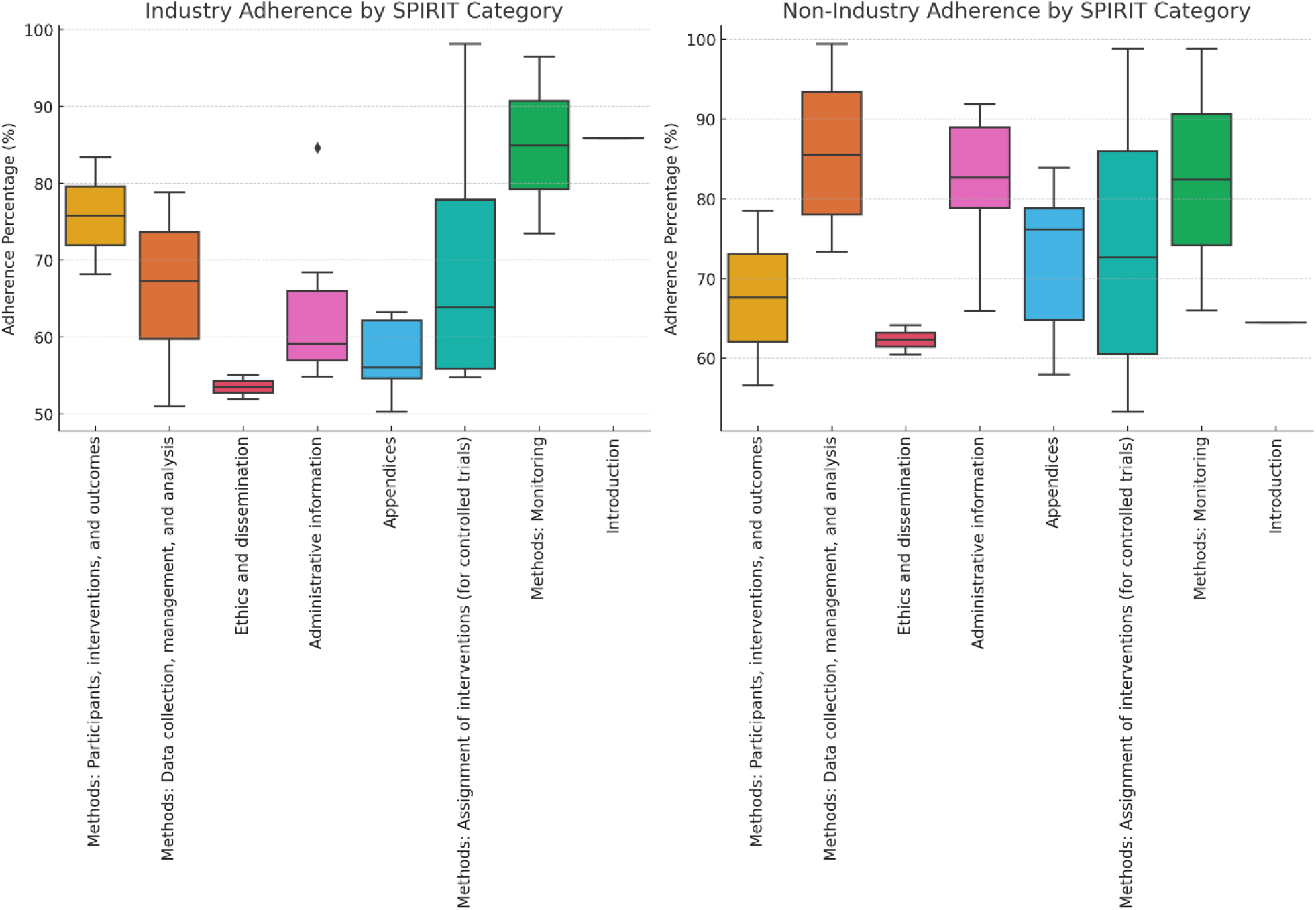
Adherence by SPIRIT Item Category

The components slightly above these, such as “Data entry and coding”, “Name and contact details of sponsor”, and “Research question described and justified”, demonstrate an adherence of approximately 42.86% for industry sponsorship and 50% for non-industry sponsorship, yielding an overall adherence of approximately 46.43%. Other SPIRIT components listed demonstrate equal adherence from both industry and non-industry sponsorships at 50%.

### 4.4 Industry vs Non-Industry Adherence

The analysis of compliance percentages across different intervention categories furthermore reveals distinct patterns between industry-sponsored and non-industry-sponsored studies. Industry-sponsored studies exhibit a higher average compliance in categories like ‘DEVICE’ (62.50%) and ‘DRUG’ (79.69%), compared to non-industry-sponsored studies, which have compliance rates of 34.38% and 46.35% for these categories, respectively. This discrepancy is particularly pronounced in the ‘DRUG’ category, where the industry compliance rate is significantly higher. Conversely, for ‘BIOLOGICAL’ interventions, non-industry sponsors show slightly higher compliance (50.00%) compared to industry sponsors (44.53%). These results suggest that industry-sponsored studies tend to achieve higher compliance in certain categories, particularly where commercial interests may be stronger, such as drugs and devices, while non-industry sponsors show comparable or slightly better compliance in categories like ‘BIOLOGICAL’ and ‘OTHER.’ This trend could reflect differences in regulatory pressures, funding availability, and study priorities between industry and non-industry sponsors.

### 4.5 Adherence by SPIRIT Categories

Analyzing the boxplots for both “Industry” and “Non-Industry” funders across various SPIRIT categories provides several key insights into adherence behaviors in clinical trials. First, the variation within each category, as indicated by the spread of the boxplots, suggests differing levels of consistency in adherence practices among trials funded by both industry and non-industry sources. For instance, categories such as “Methods: Data collection, management, and analysis” and “Ethics and dissemination” often show wider interquartile ranges, indicating a variability in how rigorously these aspects are implemented or reported across different studies.

Moreover, comparing the median adherence levels between industry and non-industry funders, we can observe that some categories might be more consistently adhered to by one group over the other. For example, “Industry” funders might show higher adherence in technical and regulatory aspects such as “Data collection methods” and “Research ethics approval”, which are critical for ensuring regulatory compliance and drug approval processes. Conversely, “Non-Industry” funders, which often include academic and non-profit research organizations, may exhibit higher adherence in areas related to “Participant engagement and outcomes”, reflecting perhaps a greater focus on patient-centered research approaches.

## 5 Conclusion

The divergence observed in current intervertebral disc degeneration (IVD) trial protocols draws attention to the critical need for standardization. The identified variations, ranging from inclusion criteria to outcome measures, emphasize the importance of establishing consensus on key elements. Standardized protocols not only enhance the comparability of study outcomes but also facilitate meta-analyses, systematic reviews, and cumulative knowledge generation. A crucial implication is the encouragement of an open-access culture in sharing clinical trial protocols, promoting transparency and collaboration. By fostering a shared understanding of interventions and methodological nuances, this approach not only expedites individual studies but also contributes to the collective advancement of spinal healthcare research. The call for standardization and open-access protocols reflects a commitment to elevating the rigor, reproducibility, and impact of future investigations in the field.

## Data Availability

The datasets generated and analyzed during the current study are openly available in the Zenodo repository. The complete dataset, titled "Dataset of Clinical Trial Protocols for Intervertebral Disc Degeneration Management (2013-2024) (1.0)," can be accessed at https://doi.org/10.5281/zenodo.13645604. This collection encompasses all relevant clinical trial protocols reviewed from January 2013 to September 2024. For additional inquiries or access to supplementary materials related to this study, please contact Francis K. Chemorion at.

